# Real-time monitoring the transmission potential of COVID-19 in Singapore, March 2020

**DOI:** 10.1101/2020.02.21.20026435

**Authors:** Amna Tariq, Yiseul Lee, Kimberlyn Roosa, Seth Blumberg, Ping Yan, Stefan Ma, Gerardo Chowell

## Abstract

**Background:** As of March 31, 2020 the ongoing COVID-19 epidemic that started in China in December 2019 is now generating local transmission around the world. The geographic heterogeneity and associated intervention strategies highlight the need to monitor in real time the transmission potential of COVID-19. Singapore provides a unique case example for monitoring transmission, as there have been multiple disease clusters, yet transmission remains relatively continued.

**Methods:** Here we estimate the effective reproduction number, Rt, of COVID-19 in Singapore from the publicly available daily case series of imported and autochthonous cases by date of symptoms onset, after adjusting the local cases for reporting delays as of March 17, 2020. We also derive the reproduction number from the distribution of cluster sizes using a branching process analysis that accounts for truncation of case counts.

**Results:** The local incidence curve displays sub-exponential growth dynamics, with the reproduction number following a declining trend and reaching an estimate at 0.7 (95% CI: 0.3, 1.0) during the first transmission wave by February 14, 2020 while the overall R based on the cluster size distribution as of March 17, 2020 was estimated at 0.6 (95% CI: 0.4, 1.02). The overall mean reporting delay was estimated at 6.4 days (95% CI: 5.8, 6.9), but it was shorter among imported cases compared to local cases (mean 4.3 vs. 7.6 days, Wilcoxon test, p<0.001).

**Conclusion:** The trajectory of the reproduction number in Singapore underscores the significant effects of successful containment efforts in Singapore, but it also suggests the need to sustain social distancing and active case finding efforts to stomp out all active chains of transmission.

## Background

The ongoing COVID-19 pandemic started with a cluster of pneumonia cases of unknown etiology in Wuhan, China back in December 2019 (1, 2). The initial cases have been linked to a wet market in the city of Wuhan, pointing to an animal source of the epidemic (3). Subsequently, rapid human-to-human transmission of the disease was confirmed in January 2020, and the etiological agent was identified as severe acute respiratory syndrome-related coronavirus 2 (SARS-CoV-2) due to its genetic similarity to the SARS-CoV discovered in 2003 (4, 5). The total global case tally has reached 750,890 infections including 36,405 deaths and involving 199 countries as of March 31, 2020 (6). As the virus continues to spread in the human population, obtaining an accurate “real-time” picture of the epidemic’s trajectory is complicated by several factors including reporting delays and changes in the case definition (7, 8). Although the COVID-19 case incidence in China has substantially declined, active transmission is now occurring in multiple countries around the world (2). Epidemiological data from these countries can help to monitor transmission potential of SARS-CoV-2 in near real-time.

Outside of China, Singapore, where the first symptomatic imported case (66 years old Chinese male) was reported on January 23, 2020, has been able to maintain relatively low COVID-19 incidence levels through active case finding and strict social distancing measures. Up until March 31, 2020, Singapore has reported 926 laboratory confirmed cases, including 24 reported case importations from Wuhan China and 501 non-Wuhan related case importations (9). Imported cases include six individuals who were evacuated from China between January 30 and February 9, 2020 and multiple citizens and long term Singapore pass holders returning from Asia, Europe and North America in late March 2020 (9-11). Moreover, Singapore has reported 3 deaths as of March 31, 2020 (9). On February 4, 2020, the Ministry of Health of Singapore reported its first local cluster of COVID-19, which was linked to the Yong Thai Hang shop (12). A total of 18 clusters with 2 or more COVID-19 cases have been reported thus far. Table 1 summarizes the characteristics of the 6 largest clusters in Singapore.

**Table 1:** Characteristics of the largest COVID-19 outbreak in Singapore as of March 17, 2020.

| **Cluster name** | **Cluster location** | **Cluster size** | **Number of Imported Cases linked to the cluster** | **Number of local cases linked to the cluster** | **Number of secondary cases in the cluster** | **Reporting date for the first case linked to cluster** | **Reporting date for the last case linked to cluster** |
| --- | --- | --- | --- | --- | --- | --- | --- |
| **Yong Thai Hang cluster** | Yong Thai Hang Medical Store on Cavan Road | 9 | 0 | 9 | 1 | February 4, 2020 | February 8, 2020 |
| **Grand Hayatt cluster** | Grand Hayatt hotel in Orchard | 3 | 0 | 3 | 0 | February 6, 2020 | February 8, 2020 |
| **The Life Church and Missions and The Grace Assembly of God cluster** | The Life Church and Missions at Paya Lebar and The Grace Assembly of God Church at Tanglin and Bukit Batok | 33 | 2 | 31 | 10 | January 29, 2020 | March 9, 2020 |
| **Seletar Aerospace Heights construction cluster** | Seletar Aerospace Heights construction site | 5 | 0 | 5 | 4 | February 9, 2020 | February 15, 2020 |
| **Wizlearn Technologies cluster** | Wizlearn Technologies in Science park | 14 | 0 | 14 | 8 | February 26, 2020 | March 3, 2020 |
| **SAFRA Jurong cluster** | SAFRA Jurong restaurant | 48 | 0 | 48 | 29 | February 27, 2020 | March 16, 2002 |

Although large-scale community transmission has not been reported in Singapore, the novel coronavirus can rapidly spread in confined and crowded places, as illustrated by large clusters of COVID-19 cases linked to the Grace Assembly of God Church, the Life Church and Missions

Singapore, Wizlearn Technologies and the SAFRA Jurong cluster (13, 14). In China, substantial hospital-based transmission of SARS-CoV-2 has been reported, with approximately 3400 cases involving healthcare workers (15). This pattern aligns well with past outbreaks of SARS and MERS (16), including substantial nosocomial transmission during the 2003 SARS outbreak in Singapore (17). To minimize the risk of hospital-based transmission of SARS-CoV-2, the Ministry of Health of Singapore has restricted the movement of patients and staff across hospitals (18). Also, because multiple unlinked COVID-19 cases have been reported in the community (19) and the recognition that a substantial proportion of asymptomatic cases may be spreading the virus (20-22), strict social distancing measures have been put in place including advising the public against large social gatherings in order to mitigate the risk of community transmission (23, 24). These social distancing measures reduce the risk of onward transmission not only within Singapore, but also beyond the borders of this highly connected nation (25). A recent influx of imported cases from Asia, Europe and North America into Singapore has triggered travel bans and restrictions for travelers and citizens (9).

The reproduction number is a key threshold quantity to assess the transmission potential of an emerging disease such as COVID-19 (26, 27). It quantifies the average number of secondary cases generated per case. If the reproduction number is below 1.0, infections occur in isolated clusters as self-limited chains of transmission, and persistence of the disease would require continued undetected importations. On the other hand, reproduction numbers above 1.0 indicate sustained community transmission (16, 27). Using epidemiological data and mathematical modeling tools, we are monitoring the effective reproduction number, Rt, of SARS-CoV-2 transmission in Singapore in real-time, and here we report the evolution of Rt by March 17, 2020. Specifically, we characterize the growth profile and the effective reproduction number during the first transmission wave from the daily case series of imported and autochthonous cases by date of symptoms onset after adjusting for reporting delays, and we also derive an estimate of the overall reproduction number based on the characteristics of the clusters of COVID-19 in Singapore.

## Methods

### Data

We obtained the daily series of 247 confirmed COVID-19 cases in Singapore between January 23-March 17, 2020 from public records of the Ministry of Health, Singapore as of March 17, 2020 (9). Individual-level case details including the dates of symptoms onset, the date of reporting, and whether the case is autochthonous (local transmission) or imported are publicly available. Clusters consisting of two or more cases according to the infection source were also assembled from case descriptions obtained from field investigations conducted by the Ministry of Health, Singapore (9). Single imported cases are analyzed as clusters of size 1 whereas unlinked cases were excluded from the cluster analysis.

### Transmission clusters

As of March 17, 2020, 18 different clusters of COVID-19 cases with 2-48 cases per cluster have been reported in Singapore. A schematic diagram and characteristics of the COVID-19 clusters in Singapore are given in Figure 1 and Table 1. The geographic location of the six clusters accounting for 45.3% of the total cases is shown in Figure 2 whereas the corresponding distribution of cluster sizes is shown in Figure 3.

**Figure 1:**
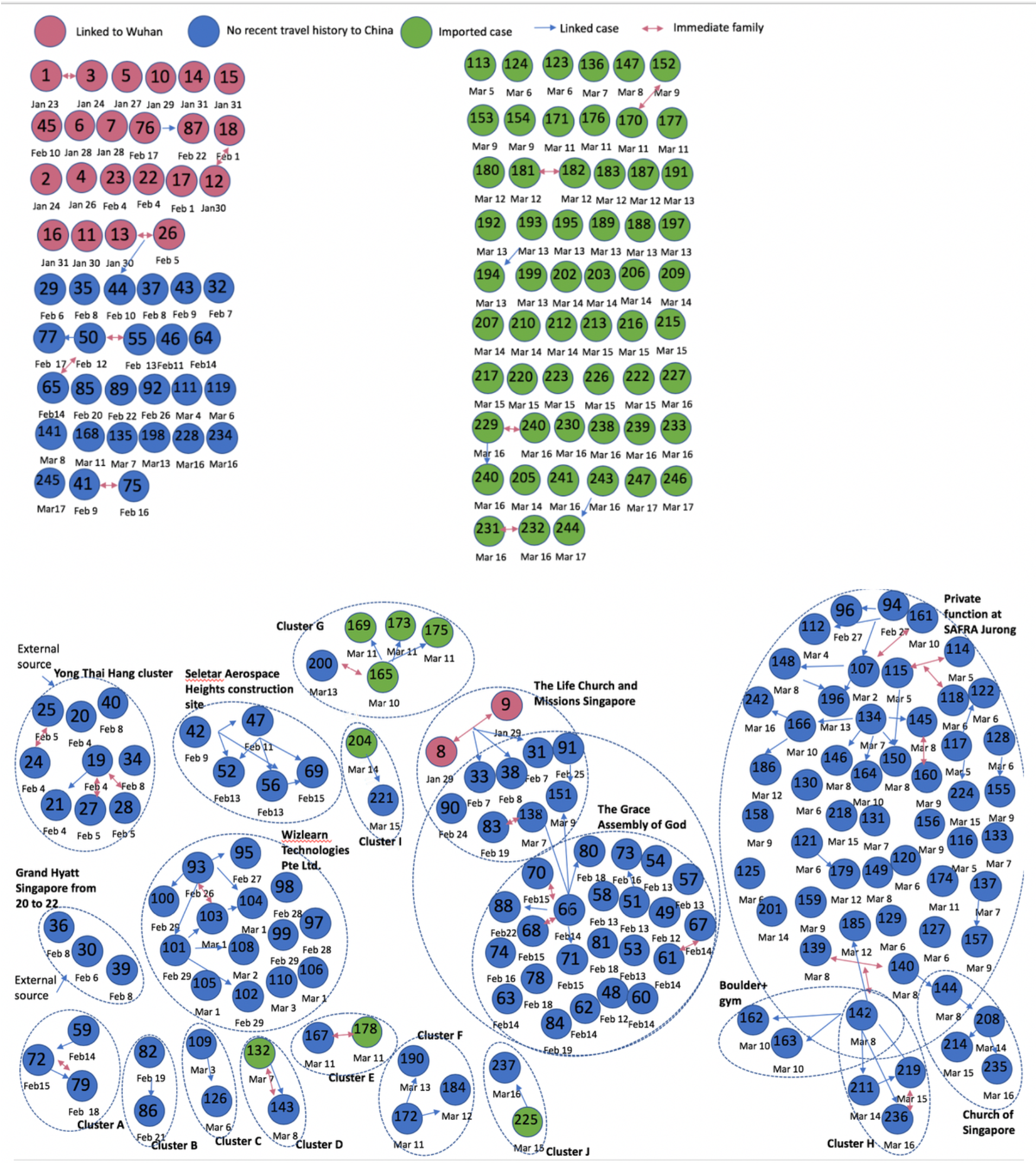
Cluster network of the cases in Singapore for the COVID-19 global pandemic as of March 17, 2020. The pink circles represent the cases linked to Wuhan, the green circles represent the non-Wuhan related case importations and the blue circles represent cases with no travel history to China. The larger dotted circles represent the COVID-19 disease clusters. Each blue arrow represents the direction in which the disease was transmitted. Pink arrows represent immediate family. Dates below the circles are the dates of case reporting.

**Figure 2:**
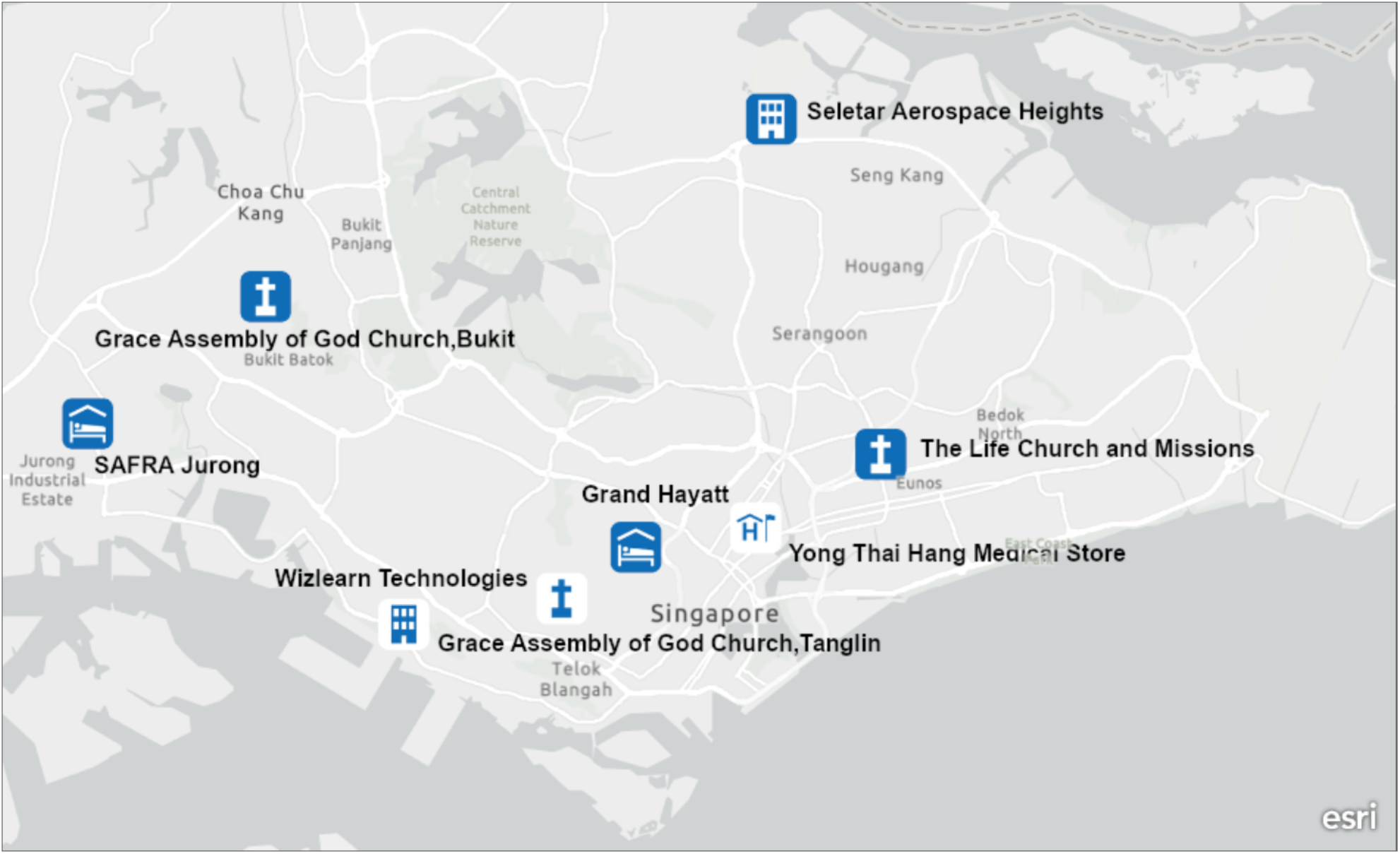
Map depicting the spatial distribution of the 6 largest COVID-19 clusters in Singapore; Grand Hayatt cluster, Yong Thai Hang cluster, Seletar Aerospace cluster, Wizlearn Technologies cluster, SAFRA Jurong cluster and The Grace Assembly of God Church and Life Church and Missions cluster as of March 17, 2020.

**Figure 3:**
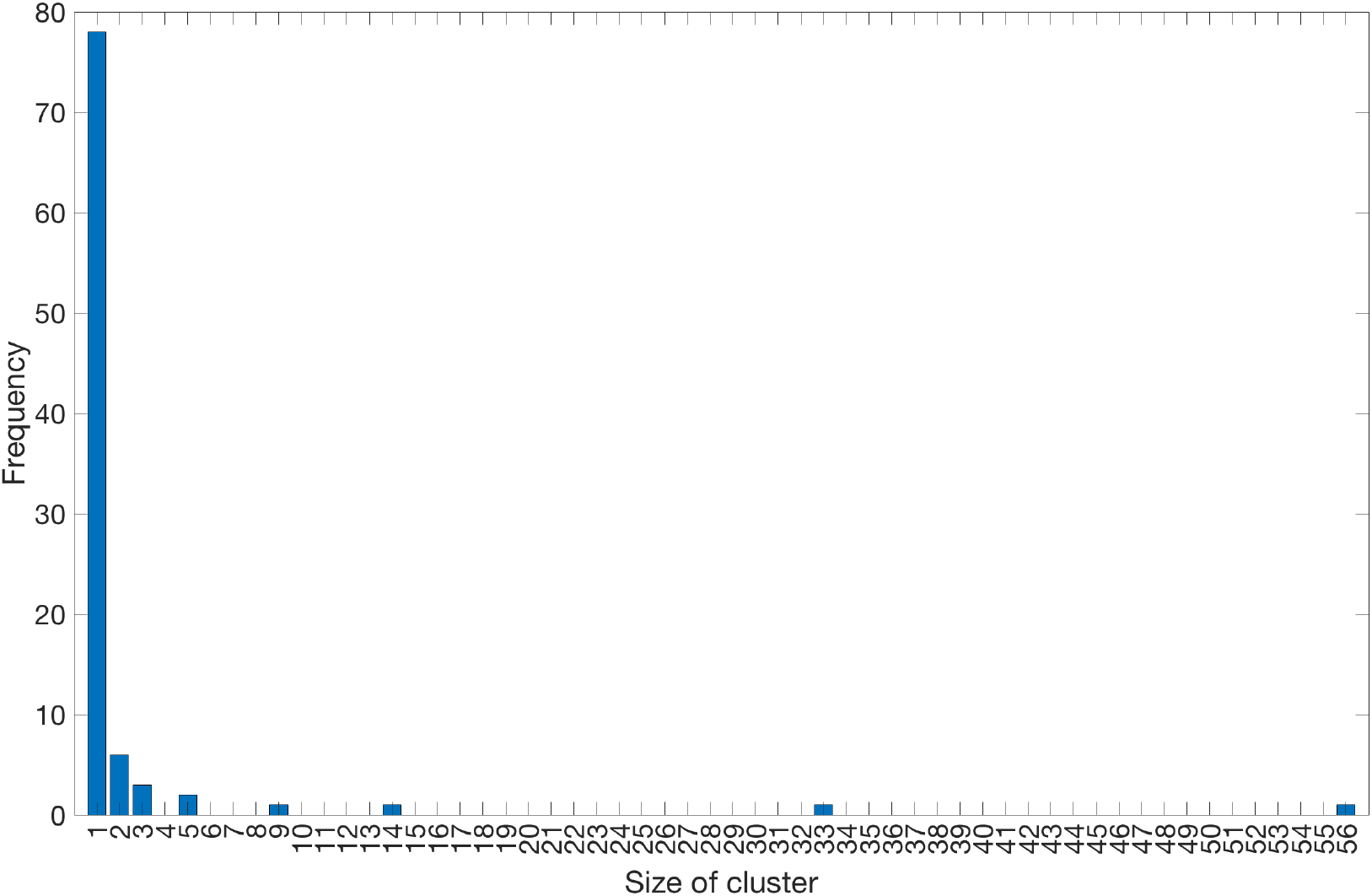
Distribution of COVID-19 cluster sizes in Singapore as of March 17, 2020.

### Yong Thai Hang cluster

This cluster with 9 cases was the first to be reported in Singapore. It has nine traceable links, including eight Chinese and one Indonesian national associated with the visit of Chinese tourists to the Yong Thai Hang health products store, a shop that primarily serves the Chinese population, on January 23, 2020. Four shop employees and the tour guide were first identified as a cluster on February 4, 2020 (12, 28, 29). The tour guide subsequently infected her husband, a newborn and the domestic helper (29). No further cases have been added to this cluster as of February 8, 2020.

### Grand Hayatt hotel

This cluster with 3 local cases was the second cluster to receive international attention, as it originated from a business meeting held at the Grand Hayatt hotel attended by Singaporean locals and the Chinese visitors from Hubei (30). Four international cases associated with this cluster had left Singapore before the onset of symptoms. All Singaporean residents associated with this cluster have recovered as of February 19, 2020 (30). No additional cases have been added to this cluster as of February 8, 2020.

### Seletar Aerospace Heights cluster

This cluster with 5 Bangladeshi work pass holders was identified on February 9, 2020. No further cases have been added to this cluster as of February 15, 2020.

### The Life Church and Missions and The Grace Assembly of God cluster

The biggest Singaporean cluster is composed of 33 cases, including two imported cases and 31 local cases. The cluster started during The Life Church and Missions service event in Paya Lebar on January 19, 2020. This event was apparently seeded by two visitors from Wuhan China who infected a couple with SARS-CoV-2 at the church. The infected couple likely passed the infection to another case during a Lunar New Year’s celebration on January 25, 2020. This case had subsequently infected Grace Assembly of God church staff at the Tanglin branch, generating secondary cases by the time he was reported on February 14, 2020. Two branches of the Grace Assembly of God church at Tanglin and Bukit Batok have been included in this cluster (28, 31). This church serves an average of 4800 people in attendance over the weekend. While the church has momentarily closed, field investigations have not led to conclusive evidence regarding super-spreading transmission (32). No further cases have been added to this cluster as of March 9, 2020.

### SAFRA Jurong cluster

The largest cluster composed of 48 local cases is linked to a private dinner function at SAFRA Jurong restaurant on February 15, 2020. The restaurant was closed for cleaning from February 16-February 19, 2020 following the dinner function. The latest case was added to this cluster on March 16, 2020.

### Wizlearn Technologies cluster

This cluster which comprises of 14 cases, was identified on February 26, 2020. Wizlearn Technologies is an e-learning solutions company. The latest case was added to this cluster on March 3, 2020.

### Church of Singapore cluster

The first case of this cluster was identified on March 14, 2020, originating as a secondary case from a case in the SAFRA Jurong cluster. This cluster is composed of 3 local cases. No further cases have been added to this cluster since March 16, 2020.

### Boulder gym cluster

The first case of this cluster was identified on March 8, 2020, also linked to the SAFRA Jurong cluster. This cluster is composed of 3 local cases. No further cases have been added to this cluster since March 10, 2020.

### Cluster A

The first case of this cluster was identified on February 14, 2020. The cluster comprise of 3 local cases. No further cases have been added in this cluster as of February 18, 2020.

### Cluster B

The first case of this cluster was identified on February 19, 2020. This cluster is composed of two local cases. No further cases have been added in this cluster since February 21, 2020.

### Cluster C

This first case of this cluster was identified on March 3, 2020. This cluster is composed of 2 local cases. No further cases have been added to this cluster since March 6, 2020.

### Cluster D

The first case of this cluster was identified on March 7, 2020. The two cases (one imported and one local) in this cluster are related to each other. No further cases have been added in this cluster since March 8, 2020.

### Cluster E

The two cases (an imported and a local case) of this cluster were identified on March 11, 2020. No further cases have been added in this cluster since March 11, 2020.

### Cluster F

The first case of this cluster was identified on March 11, 2020. This cluster is composed of 3 local cases. No further cases have been added to this cluster since March 13, 2020.

### Cluster G

This cluster is composed of 5 cases, including 4 imported cases. The first case of this cluster was identified on March 10, 2020. No further cases have been added to this cluster since March 13, 2020.

### Cluster H

The first case of this cluster was identified on March 14, 2020, a secondary case generated from a case at SAFRA Jurong cluster. This cluster is composed on 3 local cases. No further cases have been added to this cluster since March 16, 2020.

### Cluster I

The first case of this cluster was identified on March 14, 2020. This cluster is composed of one local and one imported case. No further cases have been added to this cluster since March 15, 2020.

### Cluster J

The first case of this cluster was identified on March 15, 2020. This cluster is composed of one imported and one local case. No further cases have been added to this cluster since March 16, 2020.

### Adjusting for reporting delays

As an outbreak progresses in real time, epidemiological curves can be distorted by reporting delays arising from several factors that include: (i) delays in case detection during field investigations, (ii) delays in symptom onset after infection, (iii) delays in seeking medical care, (iv) delays in diagnostics and (v) delays in processing data in surveillance systems (33). However, it is possible to generate reporting-delay adjusted incidence curves using standard statistical methods (34). Briefly, the reporting delay for a case is defined as the time lag in days between the date of onset and date of reporting. Here we adjusted the COVID-19 epidemic curve of local cases by reporting delays using a non-parametric method that employs survival analysis known as the Actuaries method for use with right truncated data, employing reverse time hazards to adjust for reporting delays as described in previous publication (35-37). The 95% prediction limits are derived according to Lawless et al. (38). For this analysis, we exclude 7 imported cases and 5 local cases for which dates of symptoms onset are unavailable.

### Effective reproduction number from case incidence

We assess the effective reproduction number over the course of the outbreak, Rt, which quantifies the temporal variation in the average number of secondary cases generated per case during the course of an outbreak after considering multiple factors including behavior changes, cultural factors, and the implementation of public health measures (16, 27, 39). Estimates of Rt>1 indicate sustained transmission; whereas, Rt <1 implies that the outbreak is slowing down and the incidence trend is declining. Hence, maintaining Rt <1 is required to bring an outbreak under control. Using the reporting delay adjusted incidence curve, we estimate the most recent estimate of Rt for COVID-19 in Singapore by characterizing the early transmission phase using a phenomenological growth model as described in previous publications (40-42). Specifically, we first characterize daily incidence of local cases for the first transmission wave (January 21-February 14, 2020) using the generalized logistic growth model (GLM) after adjusting for imported cases. This model characterizes the growth profile via three parameters: the growth rate (r), the scaling of the growth parameter (p) and the final epidemic size (K). The GLM can reproduce a range of early growth dynamics, including constant growth (p=0), sub-exponential or polynomial growth (0<p<1), and exponential growth (p=1) (43). We denote the local incidence at calendar time *t*_*i*_ by *I*_*i*_, the raw incidence of imported cases at calendar time *t*_*i*_ by *J*_*i*_, and the discretized probability distribution of the generation interval by *ρ*_*i*_. The generation interval is assumed to follow a gamma distribution with a mean of 4.41 days and a standard deviation of 3.17 days based on refs. (44, 45). Then, we can estimate the effective reproduction number by employing the renewal equation given by (40-42)

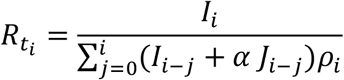

In this equation the numerator represents the new cases *I*_*i*_, and the denominator represents the total number of cases that contribute to the new cases *I*_*i*_ at time *t*_*i*_ Parameter 0≤ *α* ≤ represents the relative contribution of imported cases to the secondary disease transmission. We perform a sensitivity analyses by setting *α* = 0.15 and *α* = 1.0 (46). Next, in order to derive the uncertainty bounds around the curve of *R*_*t*_ directly from the uncertainty associated with the parameters estimates (r, p, K), we estimate *R*_*t*_ 300 simulated curves assuming a Poisson error structure (47).

### Reproduction number (R) from the analysis of cluster sizes

A second method of inferring the reproduction number applies branching process theory to cluster size data to infer the degree of transmission heterogeneity (48, 49). Simultaneous inference of heterogeneity and the reproduction number has been shown to improve the reliability of confidence intervals for the reproduction number (50). In the branching process analysis, the number of transmissions caused by each new infection is modeled as a negative binomial distribution. This is parameterized by the effective reproduction number, R, and the dispersion parameter, k. The reproduction number provides the average number of secondary cases per index case, and the dispersion parameter varies inversely with the heterogeneity of the infectious disease. In this parameterization, a lower dispersion parameter indicates higher transmission heterogeneity.

Branching process theory provides an analytic representation of the size distribution of cluster sizes as a function of R, k and the number of primary infections in a cluster (as represented in equation of 6 of the supplement of (51)). This permits direct inference of the maximum likelihood estimate and confidence interval for R and k. In this manuscript, we modify the calculation of the likelihood of a cluster size to account for the possibility that truncation of case counts at a specific time point (i.e. March 17, 2020) may result in some infections being unobserved. This is accomplished by denoting *x* as the sum of the observed number of serial intervals in a cluster. Then the likelihood that an observed cluster of size *j* containing *m* imported cases is generated by *x* infectious intervals is given by:

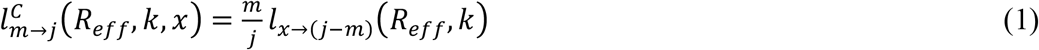

Where the likelihood of a i infections causing j infections is given by:

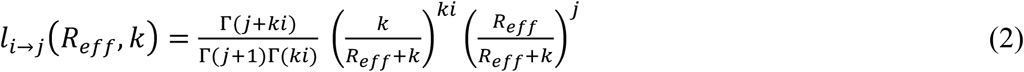

Where Γ is the gamma function.

To determine the number of observed serial intervals we observe in each cluster, we first estimate the cumulative probability distribution of the serial interval. We assume the serial interval is a gamma distribution, with a mean of 4.7 days and a standard deviation of 2.9 days (44). This translates to a shape parameter of 2.63 and a scale parameter of 1.79. We then use the difference between the onset data and the end of our study (March 17, 2020) to determine how much of the infectious period was observed. For cases that only have a report date, but no onset date, we assume an onset date that is six days earlier than the reporting date. This is based on the average duration between onset date and report date that was observed in the data. When applied to the case series, we are able to assign a total size, the number of imported cases and the observed number of infectious periods for each cluster in the case series. When no imported cases are known to be in a cluster, we assign the number of imported cases to be one as the cluster must have been initiated by someone (e.g. the index case had contact with a foreign visitor).

When equation 1 is applied to the table of cluster size characteristics, the likelihood of the data can be calculated as a function of R and k. Minimizing the likelihood produces the maximum likelihood estimates of R and k. Applying the likelihood ratio test by profiling and R and k, produces confidence intervals (52). Code was run in R version 3.6.1.

## Results

### Incidence data and reporting delays

The COVID-19 epidemic curve by the date of reporting, stratified for local and imported incidence case counts is shown in Figure 4. It shows that the majority of the imported cases are concentrated at the beginning of the outbreak (January 23, 2020 - February 3, 2020) and after March 10, 2020 in Singapore, with an average of ∼12 new cases reported per day between March 1, 2020 and March 17, 2020 (Figure 4). Out of 88 imported cases, only 14 cases have been linked to secondary cases. Meanwhile, a total of 159 autochthonous cases have been reported as of March 17, 2020 including 27 cases that are unlinked to any known transmission chains.

**Figure 4:**
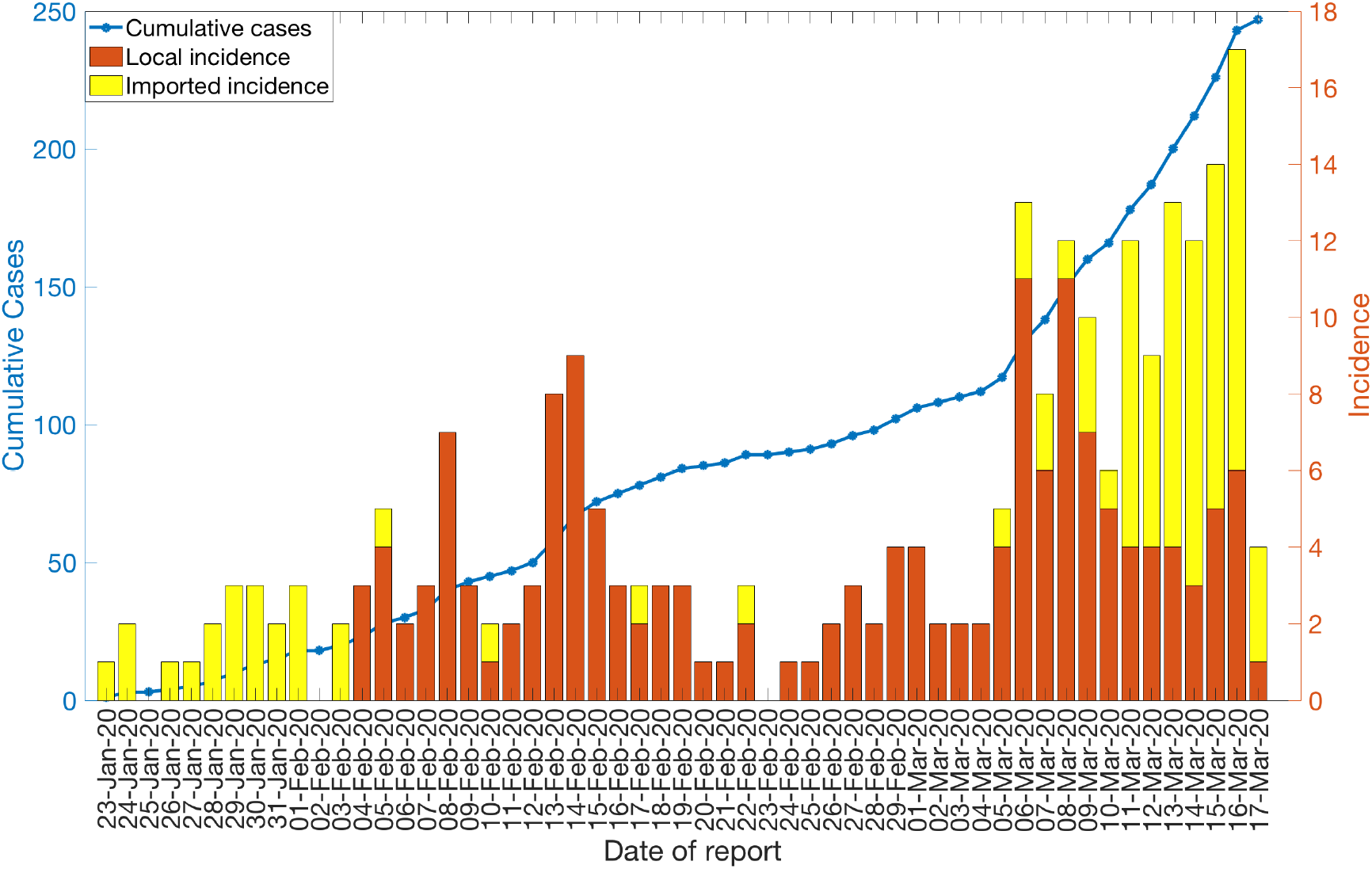
Local and imported incidence cases by date of reporting as of March 17, 2020. The solid blue line represents the cumulative case count for the COVID-19 cases in Singapore.

The reporting-delay adjusted epidemic curve of local cases by date of symptoms onset roughly displays two small waves of transmission reflecting the occurrence of asynchronous case clusters (Figure 5). Moreover, the gamma distribution provided a reasonable fit to the distribution of reporting delays for all cases, with a mean reporting delay at 6.4 days (95% CI: 5.8, 6.9) (Figure 6).We also found that imported cases tend to have shorter reporting delays compared to local cases (mean 4.3 vs. 7.6 days, Wilcoxon test, p<0.001), as imported cases tend to be identified more quickly. The mean of reporting delays for the six large clusters ranged from 4.8-13.6 days (Figure 7).

**Figure 5:**
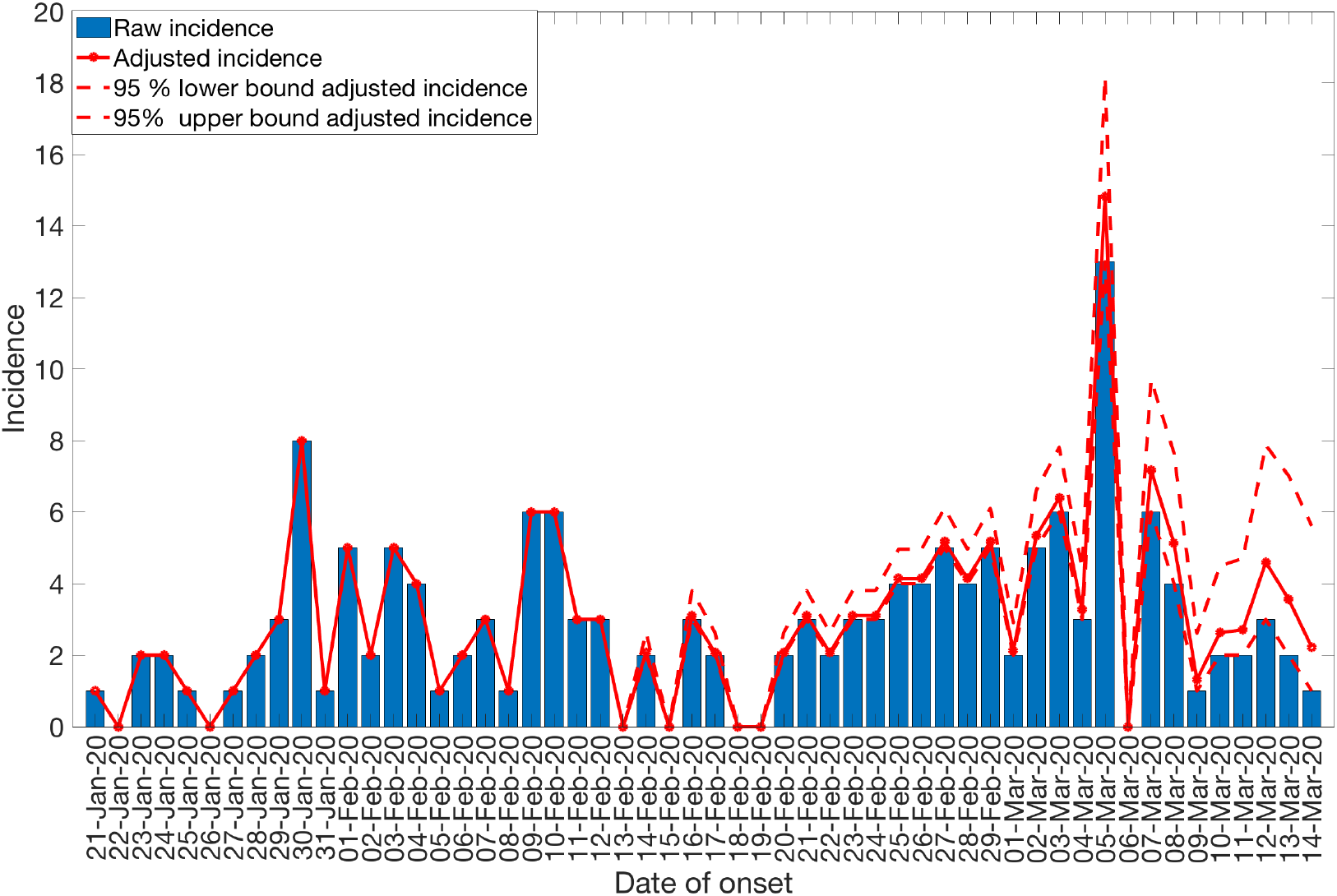
Reporting delay adjusted local incidence for the COVID-19 outbreak in Singapore as of March 17, 2020. Blue bars represent the raw incidence, red solid line represents the adjusted incidence, red dotted lines represent the 95% lower and upper bound of the adjusted incidence.

**Figure 6:**
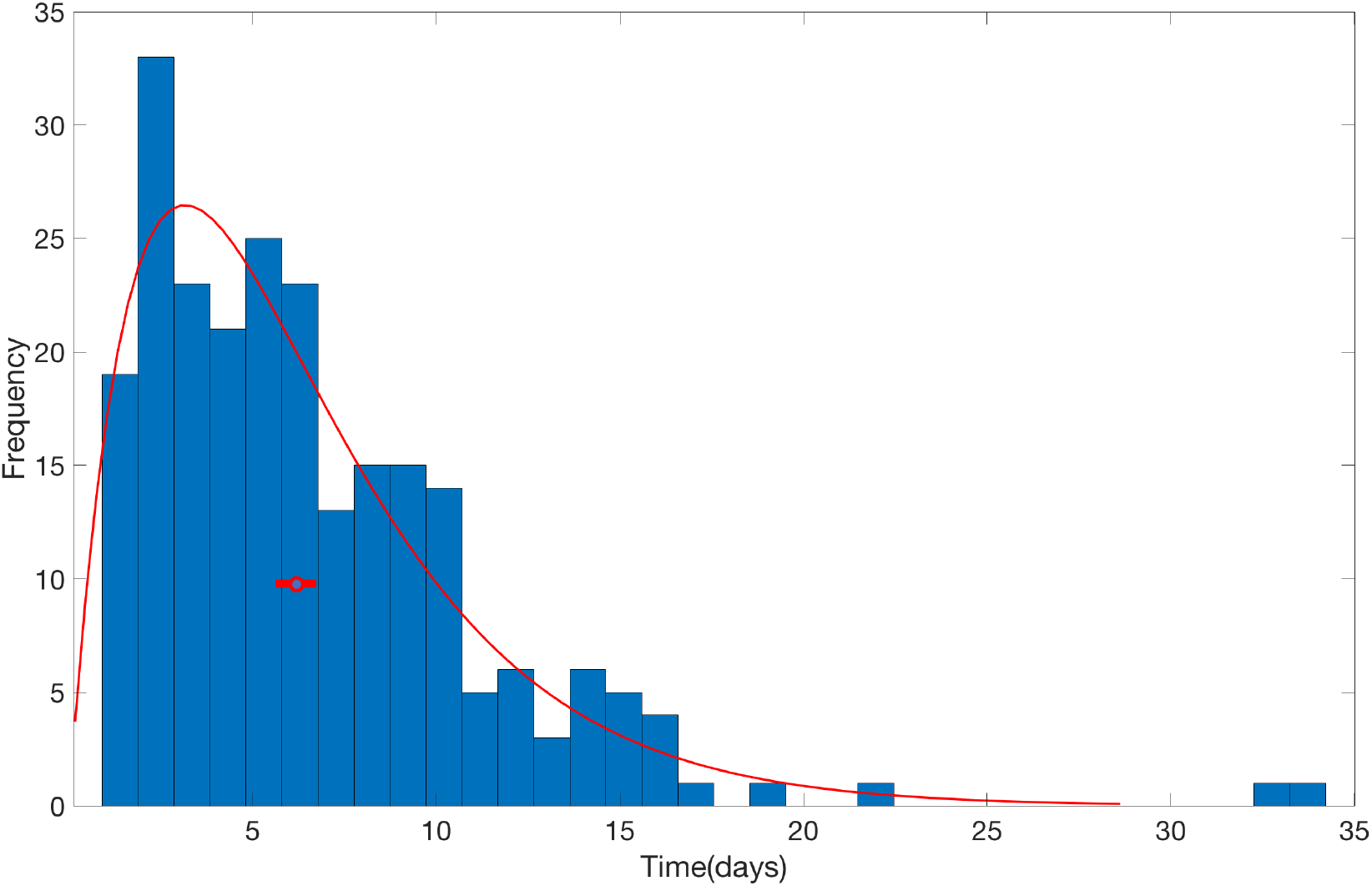
The distribution of reporting delays for all cases as of March 17, 2020. The red line represents the fit of a gamma distribution to the data. The red circle represents the mean of gamma distribution and the horizontal line represents the 95% CI.

**Figure 7:**
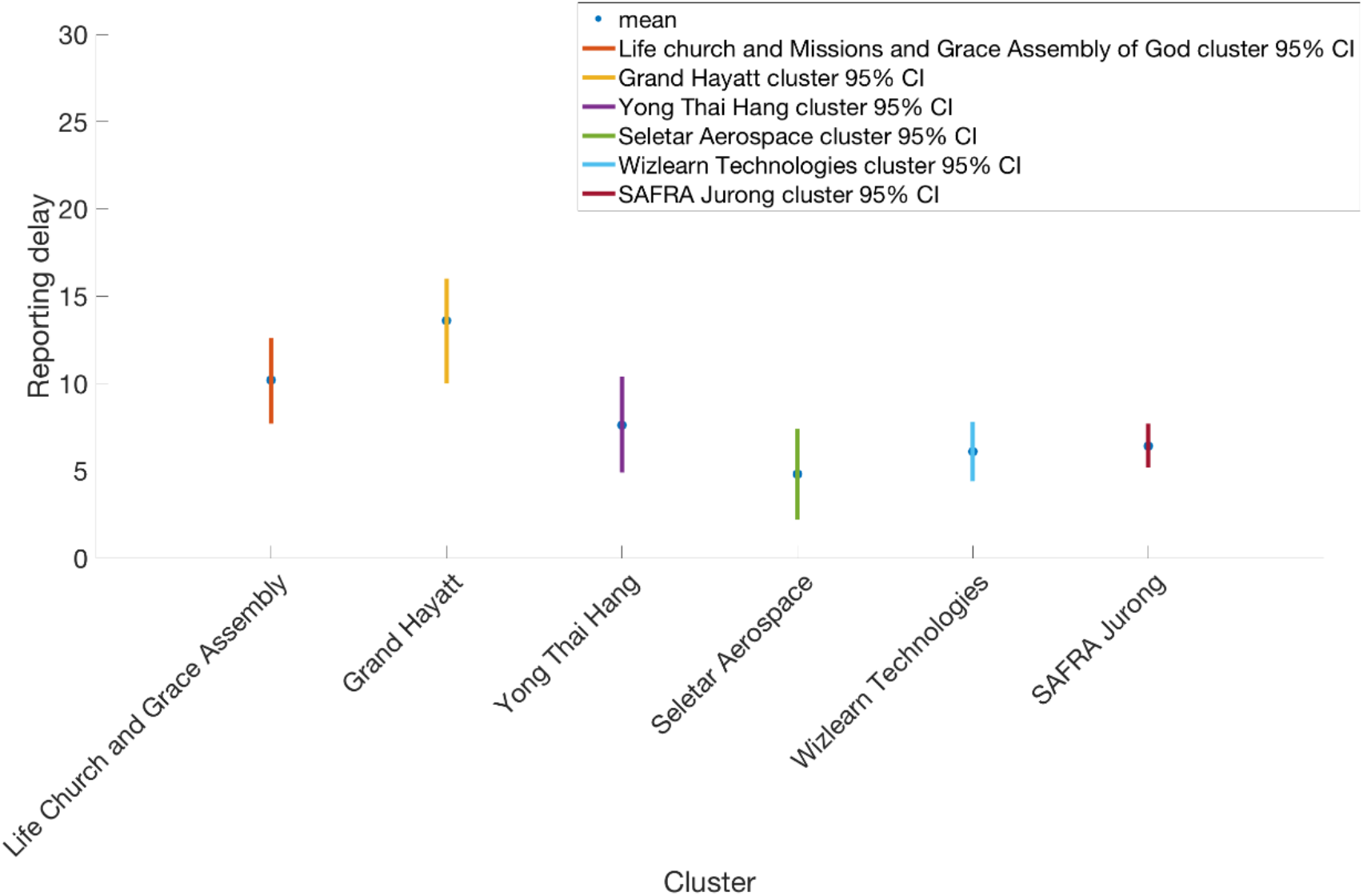
Reporting delay distribution with mean (blue circle) and 95% CI (vertical lines) for each big cluster in Singapore; Grand Hayatt cluster, Yong Thai Hang cluster, Seletar Aerospace cluster, Wizlearn Technologies cluster, SAFRA Jurong cluster and The Grace Assembly of God Church and Life Church and Missions cluster as of March 17, 2020.

### Reproduction Numbers

For the first small wave of transmission comprising the first 25 epidemic days, the delay-adjusted local incidence curve displays sub-exponential growth dynamics with the scaling of growth parameter p at 0.7 (95% CI: 0.4, 1.0), the intrinsic growth rate r estimated at 0.6 (95% CI: 0.3, 1.1) and parameter K estimating the wave size estimated at 95 (95%CI: 56, 230). Because of the sub-exponential growth dynamics, the effective reproduction number followed a declining trend with the latest estimate at 0.7 (95% CI: 0.3, 1.0) when *α* = 0.15 (Figure 8). This estimate was not sensitive to changes in parameter *α*.

**Figure 8:**
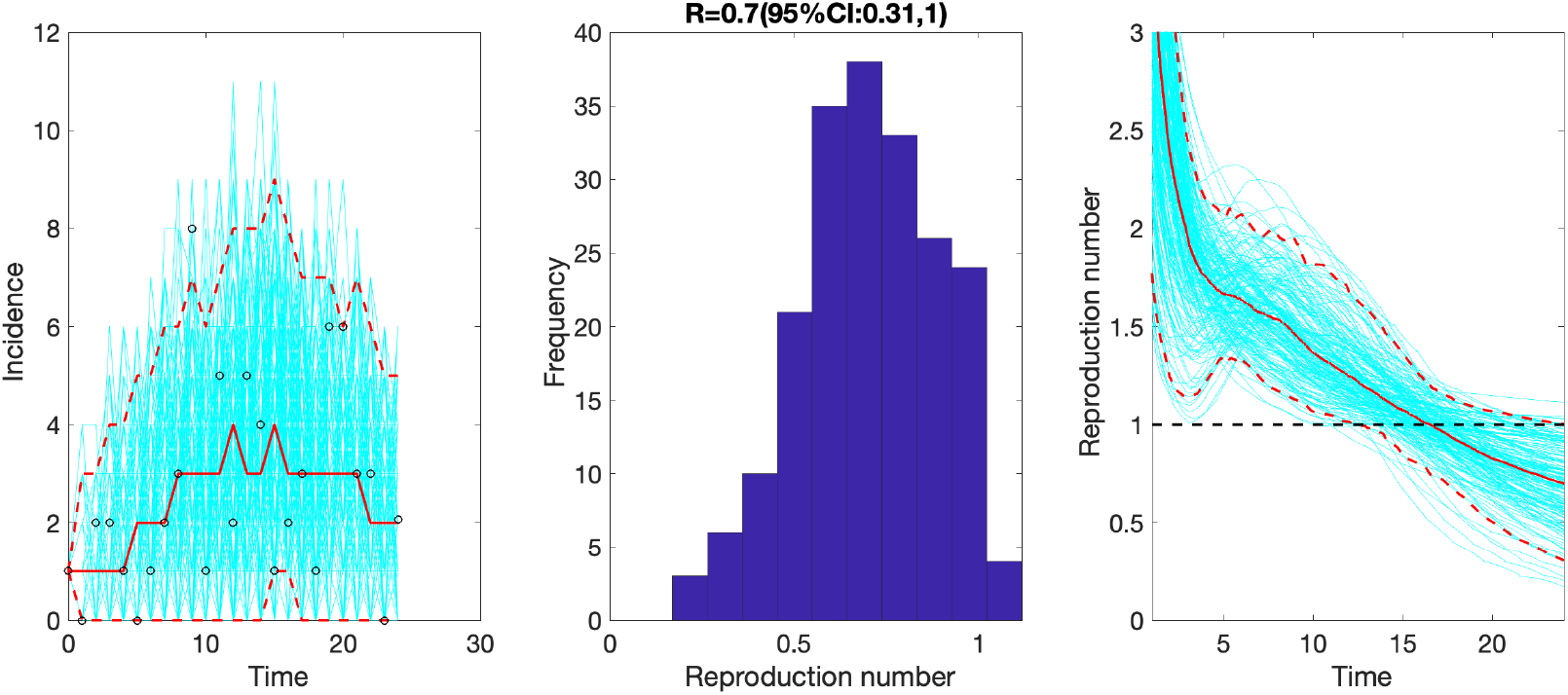
The effective reproduction number reproduction number with 95% CI estimated by adjusting for the imported cases *α* = 0.15 during the first transmission wave by February 14, 2020. The effective reproduction number followed a declining trend with the latest estimate at 0.7 (95% CI: 0.3,1.0) by February 14, 2020.

Based on the entire distribution of cluster sizes, we jointly estimated the overall reproduction number R and the dispersion parameter k as of March 17, 2020. Fitting the negative binomial distribution to the cluster data in the empirical distributions of the realizations during the early stages of the outbreak in Singapore, the reproduction number is estimated at 0.61 (95% CI: 0.39, 1.02) after adjusting for the truncation of the time series leading to the possibility that some infected cases might still cause new infections after March 17, 2020. The dispersion parameter is estimated at 0.11 (95% CI: 0.05, 0.25) consistent with SARS-CoV-2 transmission heterogeneity.

## Discussion

Overall, current estimates of transmission potential in Singapore, based on two different data sources and different methods, suggest that temporary local transmission potential of SARS-CoV-2 has occurred in Singapore while our most recent estimate of the effective reproduction number is below the epidemic threshold of 1.0 whereas the overall reproduction number derived from the distribution of cluster sizes just barely crosses 1.0 (Rt = 0.61 (95% CI: 0.39, 1.02). Temporary sustained transmission in the beginning of the epidemic can be partly attributed to multiple case importations and initiation of local transmission in the region. While large-scale local transmission has not been reported in Singapore, the fact that asymptomatic and subclinical cases are now well documented for COVID-19 (53) suggests that our estimates could be underestimated (54). On the other hand, it is not clear if asymptomatic or subclinical cases are as infectious as symptomatic cases. Indeed, we have reported that multiple local cases have yet to be traced to existing transmission chains. Additional data collected during the course of the outbreak will help obtain an improved picture of the transmission dynamics (55). These findings emphasize the need to strengthen public health interventions including active case contact tracing activities in countries with emerging transmission of SARS-CoV-2 (56). It is worth noting that imported cases have minor contribution to secondary cases in Singapore, with most of the imported cases dating back to the early phase of the epidemic and between March 10-17, 2020. However, there are examples such as the Grant Hayatt Singapore cluster and the Yong Thai Hang cluster that were linked to imported sources, and the original sources had left Singapore before these local clusters emerged (30, 57).

Our Rt estimates for Singapore are substantially lower than mean estimates reported for the COVID-19 epidemic in other parts of the world (58-66). This indicates that containment efforts have a significant impact in Singapore (Table 2). However, some differences in the reproduction numbers reported for the epidemic in China may result from different methods, differences in data sources, and time periods used to estimate the reproduction number. Similarly, a recent study has shown an average reporting delay of 6.1 days in China (67) which agrees with our mean estimate for cases in Singapore (6.4 days). Moreover, the scaling parameter for growth rate (p) indicates a sub-exponential growth pattern in Singapore, reflecting the effective isolation and control strategies in the region. This is consistent with a sub-exponential growth pattern for Chinese provinces excluding Hubei (p∼0.67), as estimated by a recent study (68); whereas, an exponential growth pattern was estimated for Hubei (p∼1.0) (68).

**Table 2:** Timeline of COVID-19 epidemic in Singapore as of March 31, 2020.

| **Date** | **Event** |
| --- | --- |
| 1/23/2020 | First imported case of SARS-CoV-2 confirmed |
| 1/23/2020-1/26/2020 | Flights to Wuhan cancelled by the Singaporean government |
| 1/29/2020 | Travelers from Hubei denied entry in Singapore |
| 2/1/2020 | New visitors with recent travel history to mainland China within the last 14 days denied entry into Singapore, or transit through Singapore |
| 2/1/2020 | Distribution of masks by the government |
| 2/4/2020 | First cases of local SARS-CoV-2 transmission |
| 2/6/2020 | First recovered patient in Singapore |
| 2/7/2020 | Singapore's outbreak response level upgraded from yellow to orange |
| 2/17/2020 | Stay at home notices issued for 14 days for all Singapore residents and long term work pass holders returning from China |
| 2/23/2020 | Travel advisory extended to visitors from South Korea |
| 2/25/2020 | Links between Grace Assembly of God cluster and The Life Church and Missions cluster established |
| 2/26/2020 | Ban on visitors arriving from Cheongdo and Daegu in South Korea. |
| 2/28/2020 | Singapore company Biotech introduced COVID-19 test kit for in-vitro case diagnosis |
| 3/4/2020 | Ban implemented on visitors arriving from South Korea, Iran and Italy |
| 3/10/2020 | 600 passengers disembarked from the Italian cruise ship, Costa Fortuna and social distancing measures announced |
| 3/12/2020 | First two deaths from COVID-19 reported |
| 3/15/2020 | Ban implemented on visitors arriving from Italy, France, Spain and Germany |
| 3/18/2020 | Announcement made for all visitors entering Singapore from March 20, 2020 onwards to observe a 14 day quarantine |
| 3/22/2020 | Ban implemented on all short term visitors arriving or transiting from Singapore from March 23, 2020 onwards |
| 3/23/2020 | Announcement made for travelers including Singapore citizens required to submit a health declaration before entering Singapore |
| 3/24/2020 | Social distancing measures reinforced including bans on large gatherings and social events |
| 3/26/2020 | Punishments announced for individuals breaching the stay at home notices |

A previous study on the 2015 MERS outbreak in South Korea reported substantial potential for superspreading transmission despite a subcritical Rt (69). The lower estimate of the dispersion parameter in our study also indicate significant transmission heterogeneity in Singapore. Super-spreading events of MERS-CoV and SARS-CoV associated with nosocomial outbreaks are well documented and driven largely by substantial diagnostic delays (16, 37). Although the average delay from onset of symptoms to diagnosis for COVID-19 patients in Singapore is at 6.4 days and no super-spreading events has been observed yet, the dispersion parameter, k<1, indicates the probability of observing large clusters and the potential for super-spreading such as the SAFRA Jurong cluster (49, 69). Therefore, public health measures enacted by public health authorities in Singapore that advise the public to avoid mass gatherings and confined places are crucial to prevent disease amplification events. However, the presence of asymptomatic cases in the community represent an ongoing threat (70, 71) although it is not currently known if subclinical cases are less infectious. This highlights the need for rapid testing of suspected cases to quickly isolate those that test positive for the novel coronavirus. To achieve this goal, public health authorities in Singapore are reactivating 900 general practitioner clinics (72). While new clusters emerge in Singapore, some clusters including the Yong Thai Hang cluster, Seletar Aerospace cluster, Wizlearn Technologies and the Grand Hayatt cluster have stabilized (no recent additional cases in most clusters). The “Grace Assembly of God and the Life Church and Missions” cluster and the “SAFRA Jurong” cluster continue to be consolidated (57, 73).

Beyond Singapore, COVID-19 cases are now being reported in 204 countries including identifiable clusters in many parts of the world (2, 74-77). Moreover, Singapore has also produced secondary chains of disease transmission beyond its borders (25). Although Singapore has been detecting and isolating cases with diligence, our findings underscore the need for continued and sustained containment efforts to prevent large-scale community transmission including nosocomial outbreaks. Overall, the current situation in Singapore highlights the need to investigate the imported, unlinked and asymptomatic cases that could be a potential source of secondary cases and amplified transmission in confined settings. Although Singapore has a world-class health system including a highly efficient contact tracing mechanism in place that has prevented to outbreak from getting out of control (25, 78), continued epidemiological investigations and active case finding efforts are needed to contain the outbreak.

Our study is not exempt from limitations. First, the outbreak is still ongoing and we continue to monitor the transmission potential of COVID-19 in Singapore. Second, onset dates are missing for twelve cases, which were excluded from our analyses. Third, we cannot rule out that additional cases will be added to existing clusters, which may lead to underestimating the reproduction number based on the cluster size distribution. Fourth, some of the cases are associated with generating secondary chains in more than one cluster, which were included in the most relevant cluster.

## Conclusion

This is a real-time study to estimate the evolving transmission potential of SARS-CoV-2 in Singapore. Our current findings point to temporary sustained transmission of SARS-CoV-2, with our most recent estimate of the effective reproduction number lying below 1.02. These estimates highlight the significant impact of containment efforts in Singapore while at the same time suggest the need to maintain social distancing and active case finding efforts to stomp out all active or incoming chains of transmission.

## Data Availability

All data are publicly available.

## List of abbreviations

COVID-19

SARS-CoV-2

## Ethics approval and consent to participate

Not applicable

## Consent for publication

Not applicable

## Conflict of Interest

The authors declare no conflicts of interest.

## Funding

G.C. is supported by NSF grants 1610429 and 1633381. G.C. and S.B. are partially supported by R01 GM 130900.

## Data declaration

All data are publicly available.

## Author Contributions

A.T, S.B., P.Y. and G.C. analyzed the data. A.T., Y. L, P.Y and S.M. retrieved and managed data;

A.T and G.C wrote the first draft of the manuscript. All authors contributed to writing and revising subsequent versions of the manuscript. All authors read and approved the final manuscript.

## Notes

### Competing Interest Statement

The authors have declared no competing interest.

